# Observational Study of Repeat Immunoadsorption (RIA) in Post-COVID ME/CFS Patients with Elevated ß2-Adrenergic Receptor Autoantibodies – an Interim Report

**DOI:** 10.1101/2023.08.31.23294813

**Authors:** Elisa Stein, Cornelia Heindrich, Kirsten Wittke, Claudia Kedor, Laura Kim, Helma Freitag, Anne Krueger, Markus Toelle, Carmen Scheibenbogen

**Affiliations:** Charité - Universitätsmedizin Berlin, corporate member of Freie Universität Berlin and Humboldt-Universität zu Berlin, Institute for Medical Immunology, Augustenburger Platz 1, 13353 Berlin, Germany; Charité - Universitätsmedizin Berlin, corporate member of Freie Universität Berlin and Humboldt-Universität zu Berlin, Dept. of Nephrology, Augustenburger Platz 1, 13353 Berlin, Germany

## Abstract

There is increasing evidence for an autoimmune aetiology in post-infectious Myalgic Encephalomyelitis/Chronic Fatigue Syndrome (ME/CFS). SARS-CoV-2 has now become the main trigger for ME/CFS. We have already conducted two small proof-of-concept studies of IgG depletion by immunoadsorption (IA) in post-infectious ME/CFS, which showed efficacy in most patients. This observational study aims to evaluate the efficacy of IA in patients with post-COVID-19 ME/CFS. The primary objective is to assess the improvement in functional ability. Due to the urgency of finding therapies for post-Covid-Syndrome (PCS), we report here the interim results of the first ten patients with seven responders defined by an increase of between 10 and 35 points in the Short-Form 36 Physical Function (SF36-PF) at week four after IA. The results of this observational study will provide the basis for patient selection for a randomised controlled trial (RTC) including sham apheresis and for a RTC combining IA with B-cell depletion therapy.

## 1. Introduction

After mild to moderate SARS-CoV-2 infection, approximately 5 - 10% of patients develop long-lasting symptoms that can be attributed to different conditions and symptom complexes referred to as Post-COVID-19 Condition or Syndrome (PCS) [1]. About half of PCS patients suffering from moderate to severe fatigue and exertion intolerance fulfil the 2003 Canadian Consensus Criteria (CCC) for Myalgic Encephalomyelitis/Chronic Fatigue Syndrome (ME/CFS) which persist beyond 20 months post-infection in most patients and encompass the full scope of post-infectious ME/CFS [2, 3]. ME/CFS can be triggered by various infections and is characterized by the core features of fatigue and exercise intolerance with post-exertional malaise (PEM) lasting at least six months after disease onset. PEM is defined as a worsening of symptoms after everyday exertion often lasting several days or longer. ME/CFS is also characterized by pain, disturbance in sleep, neurocognitive impairment, and dysregulation of cardiovascular and immune systems [4].

The mechanisms of PCS are complex and multifactorial with strong evidence for immune and vascular dysregulation similar to ME/CFS. Further, there is evidence for SARS-CoV2 persistence and clotting abnormalities in PCS [1]. Several studies described autoantibodies (AAB) to be associated with PCS including AABs to RAS (renin-aldosterone system) proteins, cytokines, antinuclear antibodies (ANA), and other AABs commonly associated with autoimmune diseases [5]. We and others found AABs to G-protein-coupled receptors (GPCR) in PCS to be associated with symptom severity. The ß2-adrenergic receptor antibody (ADRB2 AAB) was the best discriminator of PCS, and both fatigue and vasomotor symptoms were strongly associated with levels of ADRB2 AABs in PCS-ME/CFS patients [6]. These results are in line with previous findings in post-infectious ME/CFS patients, which described correlations between clinical symptoms, structural central nervous system (CNS) alterations and levels of AAB against ADRB and other GPCR [7-9].

We previously conducted a first observational study to investigate the effect of immunoadsorption (IA) in patients with infection-triggered ME/CFS with elevated ADRB2 AABs. We observed a rapid improvement of symptoms with both short and long-term responses in seven of ten patients [10, 11].

IA is an apheresis technique used to remove immunoglobulins from a patient’s plasma. Plasma is passed through an absorber that can selectively bind Immunoglobulin G (IgG) or all immunoglobulins. The absorber can be regenerated during plasma processing, allowing a highly effective removal with few side effects.

Due to the urgency of finding therapies, we here present an interim analysis of the results from the first 10 patients in our prospective observational IA study of patients with SARS-CoV-2-triggered ME/CFS. Patients with elevated ADRB2 AABs were selected based on the association of levels with symptom severity [6]. This study is performed within the Nationale Klinische Studiengruppe (NKSG) clinical trial and translational research platform for the development of treatment in PCS and ME/CFS funded by the German Ministry of Education and Research (BMBF) [12].

## 2. Materials and Methods

### 2.1. Patients

Patients were diagnosed with ME/CFS, based on the 2003 CCC for ME/CFS. Patients were recruited from October 2022 to February 2023 at the Charité Fatigue Centre at the Institute of Medical Immunology, Charité Berlin. Inclusion criteria required to have a positive PCR or antigen test for COVID-19 at disease onset and elevated ADRB2 AABs. Other relevant conditions that could cause PCS or fatigue were excluded.

### 2.2. Study Protocol

We conducted an observational study to assess the effect of IA on physical disability, symptom severity, immunoglobulin and antibody levels. The study was approved by the Ethics Committee of Charité Universitätsmedizin Berlin in accordance with the 1964 Declaration of Helsinki and its subsequent amendments. All patients provided written informed consent. Immunoadsorption was performed using TheraSorb® columns designed for the specific removal of human lambda and kappa chains including IgG (subclasses IgG1-IgG4), IgA, IgM and IgE (Miltenyi). The IA was performed within the approved use. Five sessions of IA treatments were carried out over a period of ten days with a maximum of two days in between treatments. Two further IAs will be offered to responding patients who deteriorate again.

### 2.3. Assessment of Immunoglobulins and Autoantibodies

IgG, IgA and IgM levels were measured before IA, before the 5th IA and four weeks after the first IA. Autoantibodies were measured before IA, after four IAs, and four weeks after the first IA. Antibodies against ADRB2 AABs were determined by CellTrend GmbH, Luckenwalde, Germany using ELISA technology. Pre and post-treatment samples were analyzed in the same assay run. The upper normal levels are defined based on validation studies of a healthy control group.

### 2.4. Assessment of Physical Function and Symptoms

The primary endpoint was to assess the effect of IA on physical function four weeks after IA by the Short-Form 36 version 2 questionnaire (SF 36), domain Physical Function (PF). It has been shown that an increase of at least 10 points in the SF-36 PF (range 0 - 100 = healthy) defines a clinically relevant improvement (“a little better”) and an increase of 20 points a greater clinical improvement (“much better”) [13]. Therefore, an increase of at least 10 points at four weeks after IA was defined as a minimum for a response. In addition, the Bell score, De Paul Symptom Questionnaire short form for post exertional malaise (DSQ-PEM), Fatigue Severity Scale (FSS), and weighted CCC symptoms were used to assess the presence and severity of symptoms [9, 14]. The cognitive score has been calculated as the mean of the items for memory disturbance, concentration ability and mental tiredness and the immune score as the mean of the items for painful lymph nodes, sore throat and flu-like symptoms [9]. Patient interviews were conducted before and four weeks after IA and results of the questionnaires were checked for plausibility.

### 2.5. Data Collection and Management

Study data were collected and managed using REDCap electronic data capture tools hosted at Charité - Universitätsmedizin Berlin [15, 16].

### 2.6. Statistical Analysis

Statistical data analyses were done using the software GraphPad Prism 9.5.1., © 2023 GraphPad Software. Nonparametric statistical methods were used. Continuous variables were expressed as median and interquartile range (IQR). Comparisons of different time points of two dependent groups were done using the Wilcoxon matched-paired signed-rank test. A two-tailed p-value of <0.05 was considered statistically significant.

## 3. Results

### 3.1. Patients Characteristics

All patients were diagnosed with post-COVID-19 ME/CFS with a disease duration of nine to 32 months at study inclusion. Age ranged between 33 and 59 years, six patients were female, and four patients were male. Functional disability assessed by Bell score ranged from 20 to 40, while a score of 100 represents the absence of functional disability, and the SF36-PF score ranged from five to 45 with 100 being the highest possible score. Patient characteristics are shown in Table 1.

**Table 1.** Patient characteristics and response to treatment.

| Patient No. | Gender<br>(m/f) | Age range<br>(years) | Time since COVID-19<br>(months) | Bell Score<br>before IA | SF-36 PF<br>before IA | SF-36 PF<br>4 weeks post IA | Responder<br>(yes/no) |
| --- | --- | --- | --- | --- | --- | --- | --- |
| 1 | m | 30-35 | 23 | 30 | 25 | 30 | no |
| 2 | m | 55-59 | 31 | 30 | 35 | 65 | yes |
| 3 | f | 35-40 | 10 | 20 | 15 | 25 | yes |
| 4 | f | 50-55 | 23 | 40 | 45 | 80 | yes |
| 5 | f | 55-59 | 32 | 30 | 20 | 35 | yes |
| 6 | m | 35-39 | 9 | 30 | 40 | 50 | yes |
| 7 | m | 40-45 | 25 | 30 | 25 | 70 | yes |
| 8 | f | 35-39 | 15 | 30 | 40 | 25 | no |
| 9 | f | 40-44 | 14 | 30 | 5 | 5 | no |
| 10 | f | 55-59 | 15 | 40 | 25 | 65 | yes |

IA = immunoadsorption; ME/CFS = Myalgic Encephalomyelitis/Chronic Fatigue Syndrome; SF-36-PF = Short Form-36-Physical Function

### 3.2. Course of IgG, IgA, IgM and ADRB2 AAB

In all patients, total IgG levels were within the normal range (median 11.14 g/l) prior to the first IA and decreased to median 2.29 g/l (range 0.57 – 3.23) after four days of IA. IgA and IgM were within the normal range before IA (median 1.89 g/l (IgA) and 1.19 g/l (IgM)) and decreased to median 0.48 g/l and 0.27 g/l. Four weeks after the first IA, the levels of IgG, IgA, and IgM increased again to a median of 6.64 g/l (IgG), 1.53 g/l (IgA) and 0.77 g/l /(IgM), but were still significantly lower compared to pretreatment (Fig. 1A).

**Figure 1:**
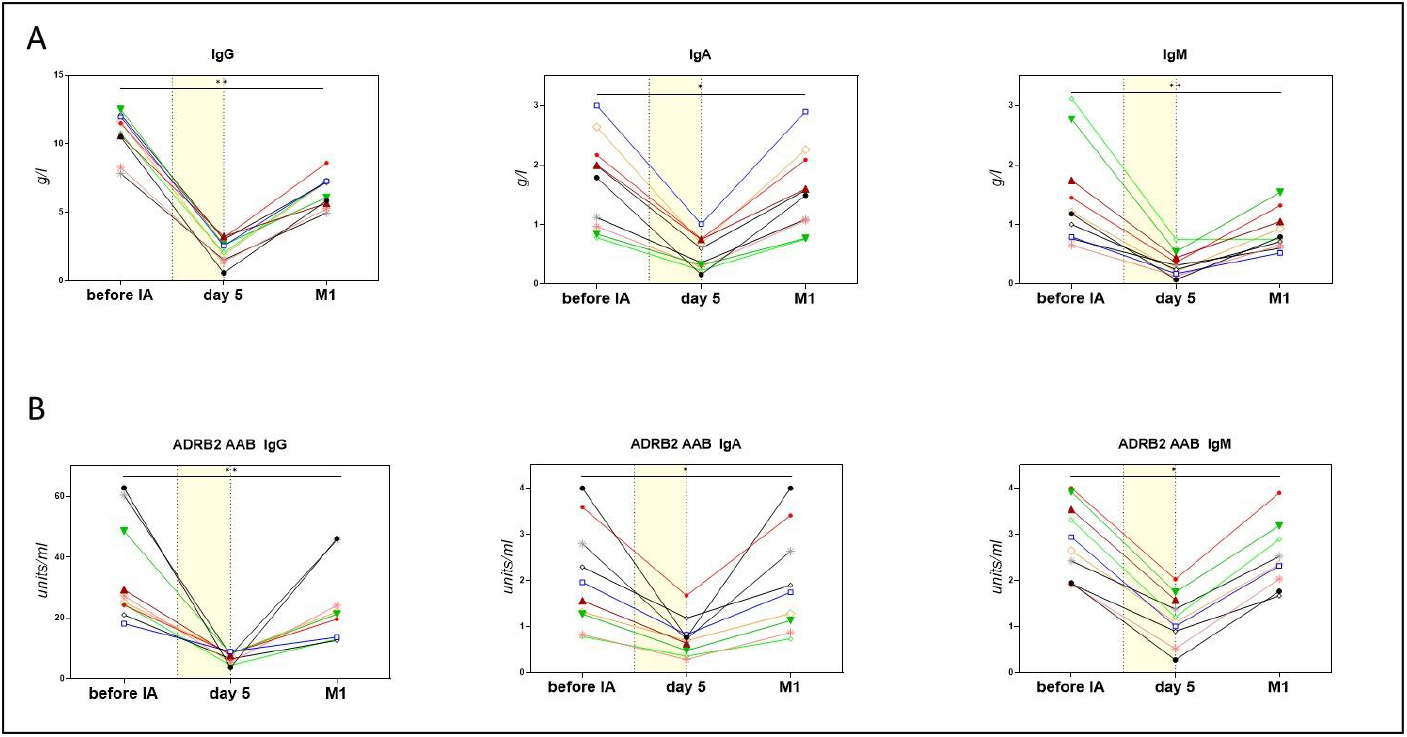
**(A) Levels of IgG, IgA, IgM** before IA, the morning before the 5th IA and 4 weeks after IA **(B) Levels of ADRB2 AAB** IgG, ADRB2 AAB IgA, ADRB2 AAB IgM before IA, the morning before the 5th IA and one month (M1) after IA. The period of IA treatment is indicated by a yellow area. ADRB2 AABs were determined by CellTrend GmbH, Luckenwalde, Germany using

ADRB2 AABs decreased in parallel with the immunoglobulin levels from a median of 26.2 U/ml (IgG)/1.8 U/ml (IgA)/2.8 U/ml (IgM) to a median of 7.7 U/ml/0.7 U/ml/1.2 U/ml after four IAs and increased again to median 21.2 U/ml (IgG)/ 1.7 U/ml (IgA)/2.4 U/ml (IgM) (Fig. 1B). We found no correlation of levels of AABs before or after treatment with response.

### 3.3. Clinical course

The SF36-PF score defined as the primary outcome parameter ranged from five to 40 at baseline (median 25). Seven patients reported an increase in SF36-PF from 10 to 35 points at week 4 after IA (to median 42.5) as shown in Table 1 and Figure 2A for all patients and Figure 2B for individual responding patients.

**Figure 2.**
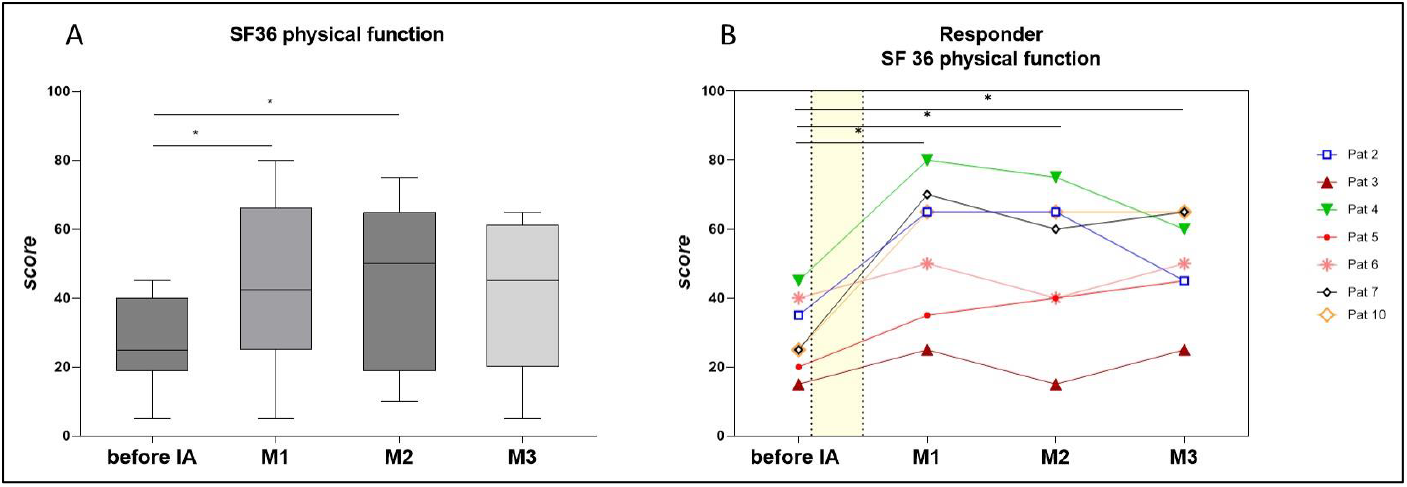
**(A) Physical function as assessed by the Short Form 36 (SF-36)** in all patients (n=10) before immunoadsorption (IA) and at months (M) 1, 2 and 3 post-IA. Statistics performed by Wilcoxon matched paired-signed rank test, * p<0,05. **(B) Physical function as assessed by the SF-36** in responders to IA (n=7) before IA and at months 1, 2 and 3 post-IA. Statistics performed by Wilcoxon matched paired-signed rank test, * p<0,05.

As shown in Figure 2B four patients reported a rapid and very good improvement in SF-36-PF with an increase of 30 – 45 points at week 4 after IA (patients 2,4,7,10). Three patients reported a minor improvement in SF-36 with 10 – 15 points (patients 3,5,6). Patient 5, however, showed a slow but steady improvement over the course of 3 months after IA from 20 before to 40 points at month 3. In two patients with a very good improvement after four weeks we observed a worsening of SF-36-PF at month 3 (patients 2 and 4).

Responders also described improvements of core symptoms pain, cognition and immunological symptoms. Figure 3A shows the course in the responding patients. Improvement of muscle pain and immune score were significant after four weeks (p<0.05), while the improvement of cognitive score and headache was not statistically significant. However, only four of seven responders reported headaches before IA.

**Figure 3:**
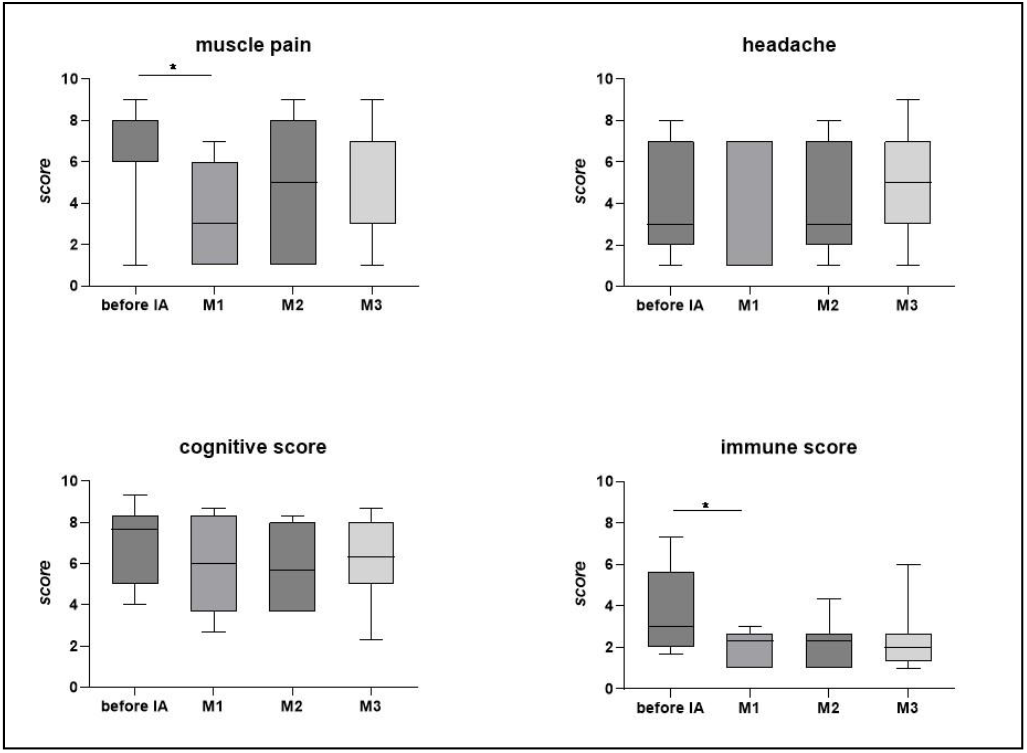
**Course of symptoms in responding patients (n=7):** muscle pain, headache, cognitive score and immune score, as assessed by weighted Canadian Consensus Criteria symptoms. Statistics performed by Wilcoxon matched paired-signed rank test, * p<0,05.

Several patients reported an initial worsening of the fatigue alongside with PEM, which they attributed to the overall strenuous process of the IA treatment. Fatigue scores assessed by the FSS showed no significant change (Fig. 4).

**Figure 4:**
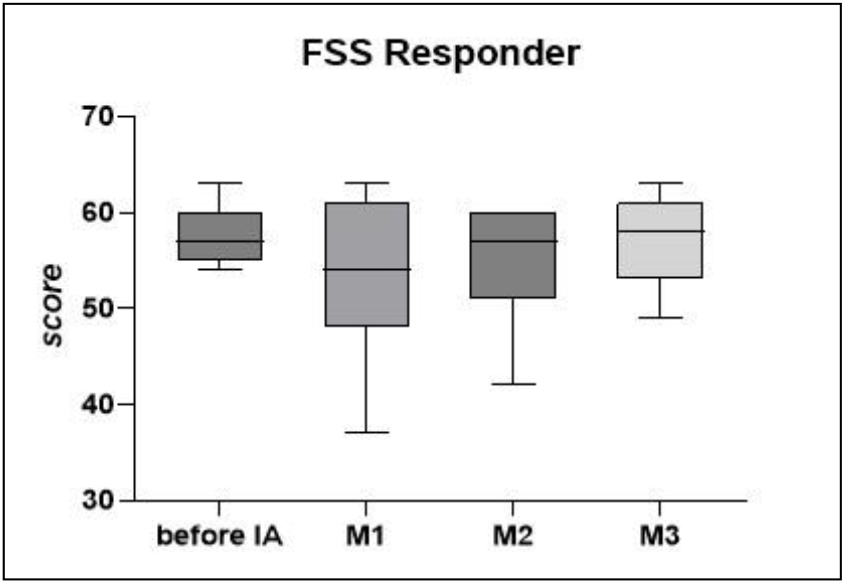
**Fatigue in responding patients (n=7)** assessed by the Fatigue Severity Scale (FSS) before IA and after months 1,2 and 3. Statistics performed by Wilcoxon matched paired-signed rank test, * p<0,05.

#### Feasibility of IA

The scheduled therapy of 5 days of IA within 10 days could be carried out in all patients in an outpatient setting and took between 4.5 to 9 hours. It could be performed with a peripheral venous catheter in six of 10 patients, four patients needed a central catheter. The IA treatment and daily travelling were rather stressful and several patients reported triggering of PEM during the therapy. To ameliorate the procedure we paid attention to good hydration and minimization of physical and mental stress as far as possible. Lorazepam for up to three days was offered as a supportive therapy.

## 4. Discussion

This interim report on the first 10 patients enrolled in our IA observational study provides first evidence that IA can improve physical function and symptoms in a subset of patients with ME/CFS following SARS-CoV2 infection. PCS is a complex condition with immune and non-immune mechanisms, thus it was important for us to understand if IA can be efficacious in SARS-CoV2 -triggered ME/CFS, before including these patients in a randomised sham-controlled trial. The SF-36-PF has been commonly used as a primary endpoint in ME/CFS clinical trials and was found to be suitable to assess the efficacy of IA in this observational study, too.

AAB levels significantly decreased after IA in all patients, both responders and non-responders and increased again after 4 weeks. There was no correlation between AAB levels and efficacy, and patients with symptom improvement at week four showed similar recurrence of AAB levels. Thus, mechanisms other than mere AAB depletion most likely account for improvement in a subset of patients. Among these is the apoptosis of AAB-producing B cells. B cell phenotyping in our previous study provided first evidence for an effect of IA on memory B cells [10]. GPCR AABs belong to a network of natural AABs that communicate with receptors regulating physiological processes in healthy individuals. There is a growing understanding of the role of these AABs in both physiological and pathophysiological processes ranging from autoimmunity to protective roles against the development of immune-mediated diseases [17]. In the case of ADRB2 AABs, we could show that they have an agonistic function in healthy individuals which is attenuated in ME/CFS [18]. Dysfunctional ADRB2 AABs were shown to be associated with Raynaud’s symptom in PCS and with brain alteration suggestive of hypoperfusion [8, 18]. Therefore, it is tempting to speculate that infection may have triggered dysfunctional GPCR AAB, which disturb receptor function. However, numerous other AABs were shown to be triggered by COVID-19 and we have no evidence that depletion of ADRB2 AABs plays a role in clinical response. Besides autoimmunity, other pathomechanisms may play a role in PCS including viral persistence, inflammation, endothelial damage or micoclotting [18]. The detailed investigation of these pathomechanisms is the subject of a comprehensive biomarker study accompanying this clinical trial within the NKSG platform [12].

## 5. Conclusion

Taken together, first data from our study provides evidence that IA has efficacy in a subset of patients and thus AABs play an important role in the pathomechanism of SARS-CoV2-triggered ME/CFS. Limitations of our study is the low number of patients completing therapy so far and the non-controlled treatment. These results are, however, the basis for recruiting patients with SARS-CoV2-triggered ME/CFS into an IA RCT with sham apheresis and a RCT combining IA with consecutive B-cell depletion. Further repeat IA will be performed in this observational trial in responding patients which deteriorate again to learn if this can lead to longer remission.

## Author Contributions

Conceptualization, C.S. and M.T.; methodology, C.S.; software, C.H. and H.F.; validation, C.S., E.S. and C.H.; formal analysis, C.H. and H.F.; investigation, E.S.; resources, A.K. and C.S.; data curation, C.H. and E.S.; writing—original draft preparation, E.S.; writing—review and editing, C.S., C.K. and L.K.; visualization, C.H.; supervision, C.S. and K.W.; project administration, E.S.; funding acquisition, C.S. All authors have read and agreed to the published version of the manuscript.

## Institutional Review Board Statement

The study was conducted in accordance with the Declaration of Helsinki, and approved by the Ethics Committe) of Charité Universitätsmedizin Berlin. Protocol code: EA2/134/22, date of approval: 28/07/2022.

## Informed Consent Statement

Informed consent was obtained from all subjects involved in the study

## Data Availability Statement

The data presented in this study will be available on request from the corresponding author after completion of the study. Due to the sensitive nature of the data and the ongoing data collection and analysis the data are not publicly available yet.

## Acknowledgments

We thank Silvia Thiel for patient care, organisation, and data management. We thank the staff of the Department of Nephrology, Charité Campus Virchow Klinikum (CVK) for patient care, organisation and performance of the immunoadsorption. We thank Harald Heidecke, CellTrend GmbH, for analysing AAB to ADRB2. We thank all patients who participated in this study and gave their consent to publish their data.

